# COVID-19 in Iran: A Deeper Look Into The Future

**DOI:** 10.1101/2020.04.24.20078477

**Authors:** Rahele Kafieh, Roya Arian, Narges Saeedizadeh, Shervin Minaee, Zahra Amini, Sunil Kumar Yadav, Atefeh Vaezi, Nima Rezaei, Shaghayegh Haghjooy Javanmard

## Abstract

The novel corona-virus (COVID-19) has led to a pandemic, affecting almost all countries and regions in a few weeks, and therefore a global plan is needed to overcome this battle. Iran has been among the first few countries that has been affected severely, after China, which forced the government to put some restriction and enforce social distancing in majority of the country. In less than 2 months, Iran has more than 80,000 confirmed cases, and more than 5,000 death. Based on the official statistics from Iran’s government, the number of daily cases has started to go down recently, but many people believe if the lockdown is lifted without proper social distancing enforcement, there is a possibility for a second wave of COVID-19 cases. In this work, we analyze at the data for the number cases in Iran in the past few weeks, and train a predictive model to estimate the possible future trends for the number of cases in Iran, depending on the government policy in the coming weeks and months. Our analysis may help political leaders and health officials to take proper action toward handling COVID-19 in the coming months.

## 1 INTRODUCTION

An outbreak of a pneumonia with unknown origin is reported in the last days of December 2019 in Wuhan, China [1]. The World Health Organization named this disease COVID-19 after that genetic sequencing revealed the same origin of the etiologic agent with coronaviruses [2].The mean incubation period of this virus is estimated to be 6.4 days (2-14 days) and the infected patient is asymptomatic in the incubation period [3, 4]. Patients infected with this virus have flu-like symptoms, including fever, cough, fatigue and dyspnea [5]. The overall death rate is estimated to be 2.3% but is higher in elderly and those with comorbidities [6]. Although public health measures have already implemented in china, but more than 2 million people in almost all countries and territories are infected during a 4-month period [7].

One of the most important concerns in dealing with the influenza-like illness (ILI) pandemics such as COVID-19 is early identification and short-term (online) estimation of its final size and peak time. This early prediction using mathematical models and combining with a small amount of existing data would effectively help the governments and public health officials to put in place appropriate prevention and control strategies. For answering this issue many mathematical models are already used for prediction ILI pandemics. Two approaches are generally considered in the literature on forecasting ILI pandemics. The first is focused on short-term estimation, and the others try to predict a long-term estimation. Based on some prominent studies in this filed [8, 9], deep learning methods conquered other classical models in short-term estimation of pandemics. In special case of COVID-19, given the novelty of the subject, most studies have already focused on short-term prediction; however, to the best of our knowledge, no work is already published on prediction of occurrence of COVID-19 using deep learning models. Limited number of works have already used deep learning in diagnosis of COVID-19 using medical images [10].

In this study, we provide a deep learning based prediction method that can assist medical and governmental institutions to prepare and adjust as pandemics unfold. To this end, we develop multiple models describing epidemic and compare their performance and effective features in forecasting. To make sure our methodology is generalize-able, in addition to Iran’s data (which is our main focus in this paper), we also apply this framework to several other countries and show consistent finding for all of them. Figure 1 provides an overview on countries selected for performance evaluation in this paper.

**Figure 1:**
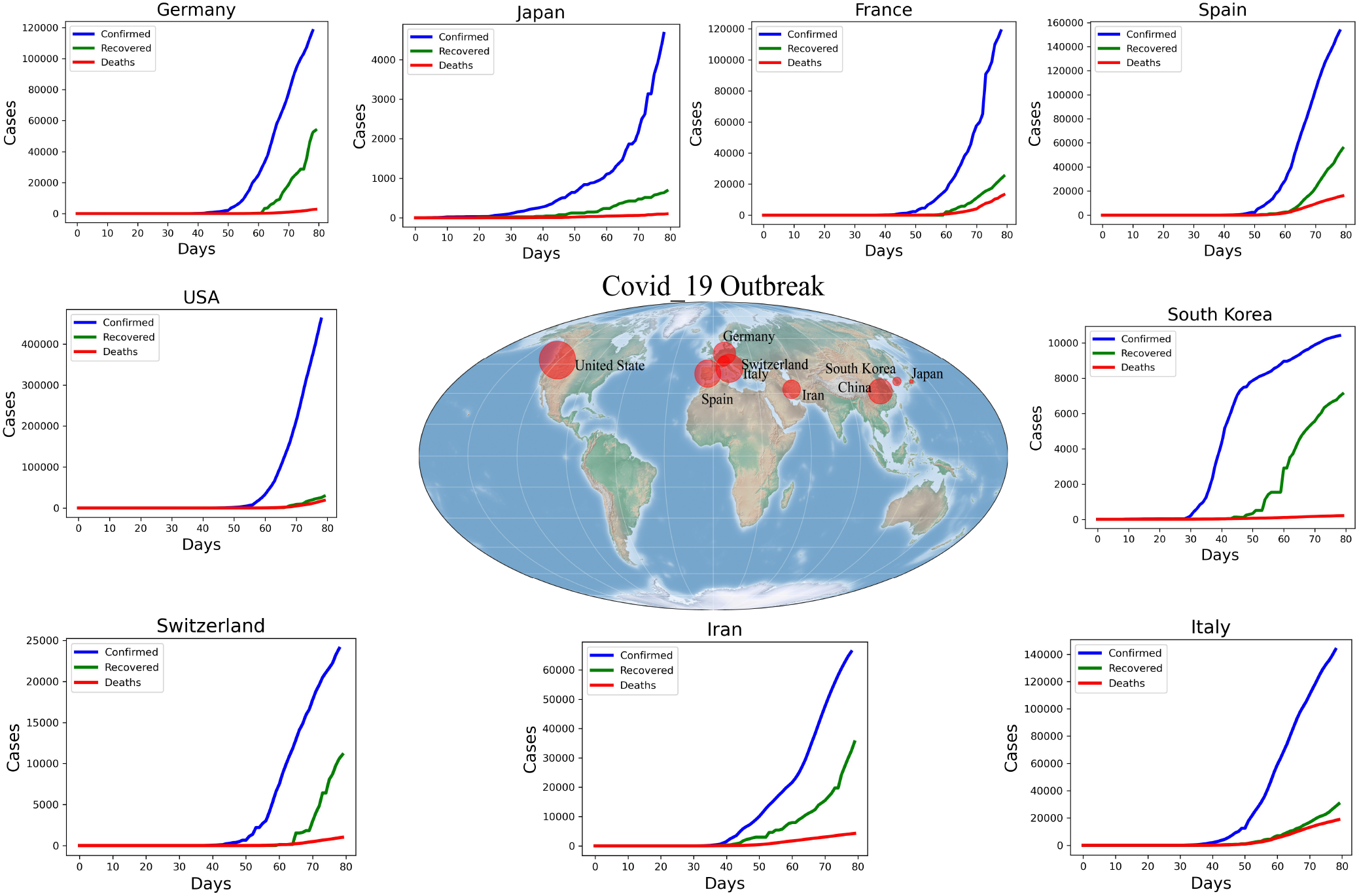
cases around the world.

The structure of the rest of this paper is as follows. Section 2 provides a overview of some of the representative works which are relevant to this work. Section 3 presents the details of the data sources used in this study. Section 4 gives a quick introduction to the machine learning algorithms used for training a predictive model. Section 5 provides a detailed quantitative and qualitative analysis of the accuracy of our forecasting model, and some of the possible future trends for COVID-19 situation in Iran and other countries. Finally the paper is concluded in section 6 by providing a discussion on the current situation and the future of COVID-19 in Iran based on the current data.

## 2 RELATED WORKS

Two approaches are generally considered in the literature on fore-casting ILI pandemics. The first is focused on short-term estimation, and the others try to predict a long-term estimation. Based on some prominent studies in this filed [8, 9], deep learning methods conquered the other competitors in short-term estimation of pandemics. In special case of COVID-19, given the novelty of the subject, most studies have already focused on short-term prediction; however, to the best of our knowledge, no work is already published on prediction of occurrence of COVID-19 using deep learning models. Here we go over some of the representative ones.

In [11], Fan et al. investigated the effect of early recommended or mandatory measures on the reducing the crowd infection percentage, using a crowd flow model.

In [12], Hu et al. developed a modified stacked auto-encoder for modeling the transmission dynamics of the epidemics. Using this framework, they forecasted the cumulative confirmed cases of COVID-19 across China from Jan 20, 2020 to April 20, 2020.

In [13], Roosa et al. used phenomenological models that have been validated during previous outbreaks to generate and assess short-term forecasts of the cumulative number of confirmed reported cases in Hubei province, the epicenter of the epidemic, and for the overall trajectory in China, excluding the province of Hubei. They collected daily report of cumulative confirmed cases for the 2019-nCoV outbreak for each Chinese province from the National Health Commission of China. They provided 5, 10, and 15 day forecasts for five consecutive days, with quantified uncertainty based on a generalized logistic model.

In [14], Liu and colleagues used early reported case data and built a model to predict the cumulative number of cases for the COVID-19 epidemic in China. The key features of our model are the timing of implementation of major public policies restricting social movement, the identification and isolation of unreported cases, and the impact of asymptomatic infectious cases.

In [15], Kucharski et al. Combined a mathematical model of severe SARS-CoV-2 transmission with four datasets from within and outside Wuhan, and estimated how transmission in Wuhan varied between December, 2019, and February, 2020. They used these estimates to assess the potential for sustained human-to-human transmission to occur in locations outside Wuhan if cases were introduced.

In [16], Peng and colleagues analyzed the COVID-19 epidemic in China using dynamical modeling. Using the public data of National Health Commission of China from Jan. 20th to Feb. 9th, 2020, they estimated key epidemic parameters and make predictions on the inflection point and possible ending time for 5 different regions.

In [17], Remuzzi analyzed the COVID-19 situation in Italy, and mentioned if the Italian outbreak follows a similar trend as in Hubei province, China, the number of newly infected patients could start to decrease within 3–4 days, departing from the exponential trend, but stated this cannot currently be predicted because of differences between social distancing measures and the capacity to quickly build dedicated facilities in China.

In [18], Sajadi et al. tried to predict potential spread and seasonality for COVID-19 based on temperature, humidity, and latitude information. They found that the distribution of significant community outbreaks along restricted latitude, temperature, and humidity are consistent with the behavior of a seasonal respiratory virus.

## 3 RESEARCH DATA

This study uses four data sources to predict COVID-19 disease, including COVID-19 data, basic information, detailed information for each country, as well as SARS Data. The **COVID-19 Data** (by John Hopkins university) contains daily number of confirmed/ death / recovered people. The **Basic information** contains the information about date/country/province of the cases. The **detailed information for each country** (around 18576 observations) includes information such as Region/ Population/Area (sq. mi.)/ Pop. Density (per sq. mi.)/ Coastline(coast/area ratio)/ Net migration/ Infant mortality (per 1000 births)/GDP/Literacy (%)/ Phones (per 1000)/ Climate/ Birthrate/ Death-rate/ Agriculture/ Industry/ Service/Arable (%)/ Crops(%)/. The **Occurrences of SARS Data** (2539 observations) contains information on the number of confirmed/ death / recovered people for SARS.

More details on how the data is used, is as follows. We collected the COVID-19 data from John Hopkins University [19] and [20]. The dataset contains cumulative number of confirmed, death and recovered COVID-19 cases in various locations across the world for different dates. Basic information (Date, Country, Province) were also provided by John Hopkins University. We also added more detailed information for each country; this includes Region, Population, Area, etc. Detailed information for each country is according to information in [21]. Different combinations of the involved data are evaluated on proposed models and “the most effective combination” and “optimal lag parameter” are selected based of effectiveness in data modeling (section 5). In this work, we propose to show the performance of different deep learning (DL) and machine learning (ML) models for COVID-19 data prediction for above mentioned dataset. To have a successful prediction using deep learning based models, it is essential to have a large-scale dataset. However, since COVID-19 is a recent disease in the world, the available of data is limited and training a deep learning model from scratch on this dataset becomes very challenging. Therefore, with respect to intrinsic similarities of COVID-19 to pandemics like SARS, we also use a public dataset: [22] from November 2002 to July 2003 on SARS data. We first train the model with large-scale data for SARS; the weights of the trained model are then used as pre-trained network and transfer learning is then utilized by training the weights of the last fully connected layers using COVID-19 dataset.

## 4 ANALYSIS METHOD

The overall framework of the proposed model is shown in figure 2. First the relevant information are extracted and processed from data sources. Then the model is pre-trained on SARS data (since more labeled data is available on SARS). Those models are then fine-tuned (re-trained) on COVID-19 data. Finally the models’ performances are measure using mean average percentage error (MAPE) metric. We experimented with more than 8 machine learning models, but to be concise, only report the result of four promising ones, which includes, Random Forest (RF) [23], multi-layer perceptron (MLP) [24], Long short-term memory (LSTM) [25] with regular features (LSTM-R), STM with extended features (LSTM-E), and Multivariate LSTM (M-LSTM).

**Figure 2:**
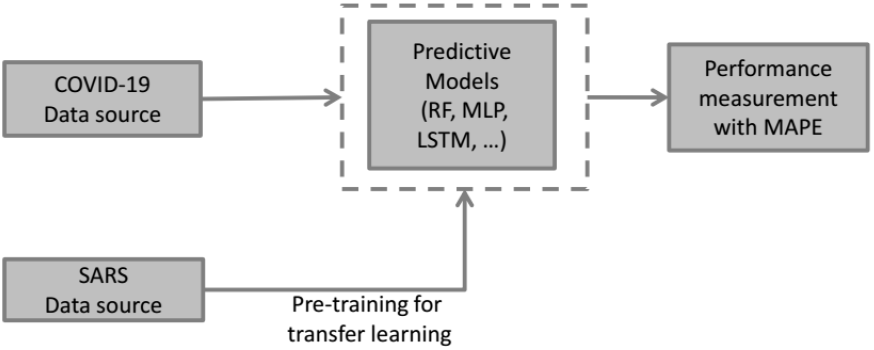
COVID-19 disease prediction models.

For each of these models, different structures (hyper-parameters and parameters) are examined and best performing architectures are summarized in Table 1.

**Table 1:** The best selected architecture for each model.

| Model | Layers | Filters |
| --- | --- | --- |
| Random Forest | n_estimators = 300 | random_state = 10 |
| MLP | 6 | 128, 128, 256, 256, 256, 1 |
| LSTM-R | 2 LSTM+ 2 Dense | 64,32,32,1 |
| LSTM-E | 2 LSTM+ 2 Dense | 64,32,32,1 |
| M-LSTM | 2 LSTM+ 2 Dense | 64,32,32,1 |

For comparison of the models, MAPE metric is used to measure the size of the error in percentage terms regarding to the actual values. MAPE is calculated using equation 1:

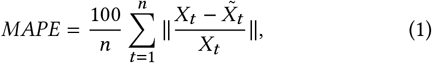

where *X*_*t*_ is the actual value and 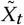 is the corresponding estimated value for *t*^*th*^ sample from all n available samples.

### 4.1 Random Forest

One of the models used in our work is Random Forest (RF). RF is essentially an ensemble of decision trees, i.e. it predicts the target value by training several decision tress and combining their results. One nice feature of RF is that it can be used for both regression and classification problems. Once the model is trained, the average predicted score of different trees can be used to predict the value of test samples. Figure 3 shows the overall diagram of a RF.

**Figure 3:**
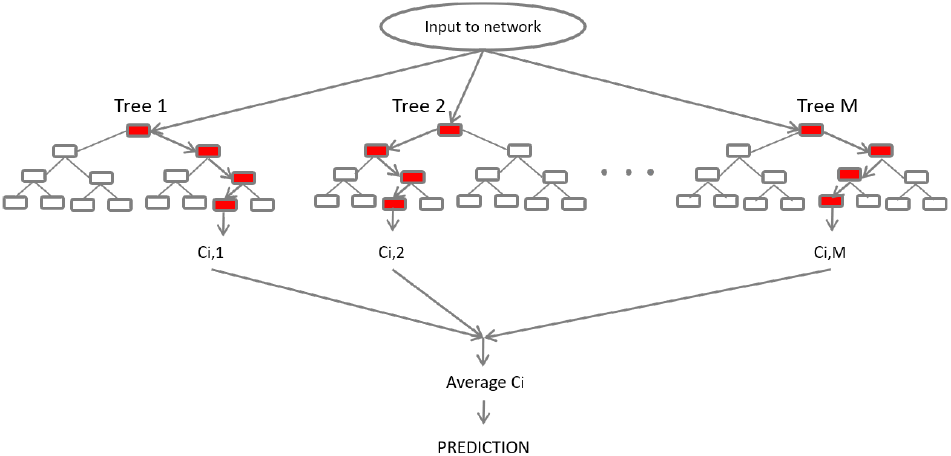
The overall diagram of a RF.

### 4.2 Multi-layer Perceptron (MLP)

Multi-layer Perceptron (MLP) (also known as feedforward network) is a popular neural network, which uses a cascade of several non-linear transformations (or layers) to make a prediction. The input features are sometimes called input layer, and the intermediate transformations are called hidden layer. All nodes in hidden layers use a nonlinear activation function (Figure 4). Figure 4 shows the overall structure of a MLP model.

**Figure 4:**
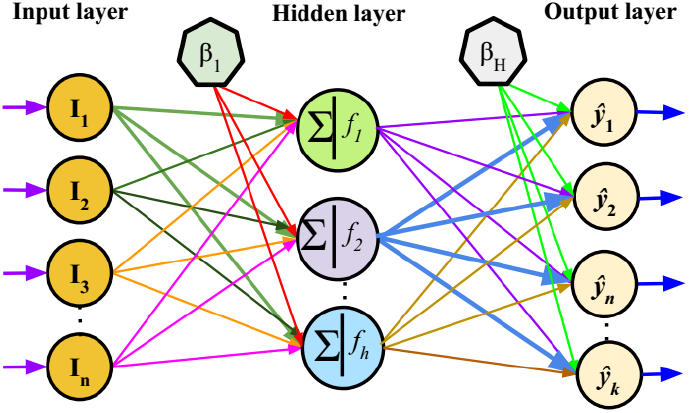
The overall structure of a neuron and a MLP model. [26].

### 4.3 Long short-term memory (LSTM)

Unlike static data (such as images), time series also adds the complexity of a sequence dependency among the input variables [25] (Figure 6), and ideally require a model with sequential processing capability. The vanilla neural networks (such as MLP) do not have the sequential processing power. However their is an extension of feed-forward neural networks for this purpose, called recurrent neural networks, where at each step the input from the current time and the hidden state from the previous time-stamp is used to make a prediction.

Figure 5 shows the architecture of RNN models. Figure 6 shows the high-level architecture of a single LSTM unit.

**Figure 5:**
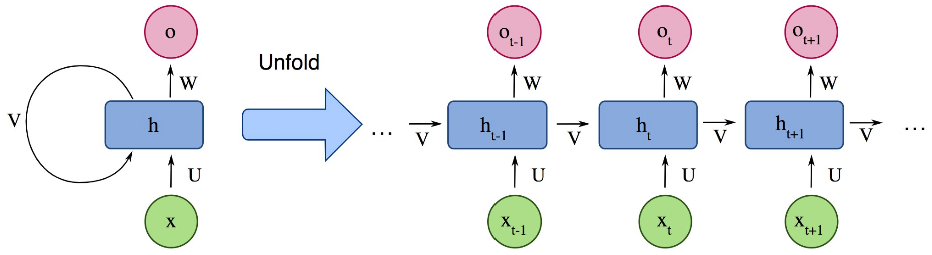
RNN model.

**Figure 6:**
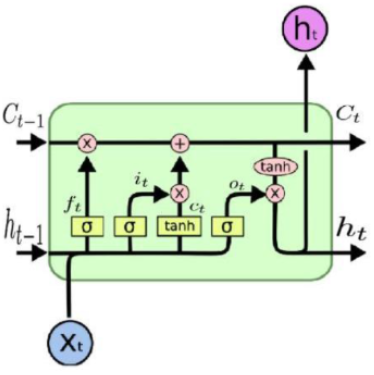
LSTM model. Courtesy of Karpathy.

#### 4.3.1 LSTM with regular features (LSTM-R)

In this application of LSTM, we originally deal with three different occurrences: number of confirmed/ death/ recovered people. Using regular features, we feed lagged samples of each occurrence to predict next values (a single-input and single-output (SISO) format)

#### 4.3.2 LSTM with extended features (LSTM-E)

By adding extended occurrences from other two types in prediction of the next value for each class (confirmed/ death / recovered), we accept a multi-input and single-output (MISO) format. We used Stacked LSTM model in section 4.3 for blocks of this network.

#### 4.3.2 Multivariate LSTM (M-LSTM)

An alternate time series problem is the case where there are multiple parallel time series and a value must be predicted for each. Now, we may consider number of occurrences in all classes (confirmed/ death / recovered) as input data and predict the value for each of the three time series for the next time step (a multi-input and multi-output (MIMO) format).

## 5 RESULTS

The machine learning models are trained and tested based on 18576, 18576, and 17569 occurrences of daily number of confirmed, death, and recovered COVID-19 cases. A lag of six days was applied to the data. The dataset is divided into training and test data sets. (Data from 22 January, 2020 til 23 March 2020 was used as the training set and the data from 24 March 2020 till 2 April 2020 was used in test stage for performance evaluation of the proposed prediction method). The training data is further divided to train and validation subsets using ratio of 7:3 based on the dates. The networks are pre-trained with 2539 occurrences for daily number of confirmed, death, and recovered cases for SARS data. Each model is tested with many different architectures and the best performance is achieved with architectures described in Table 2. We used implementation in Keras package in the Python version 3.7.3 [27].

**Table 2:**
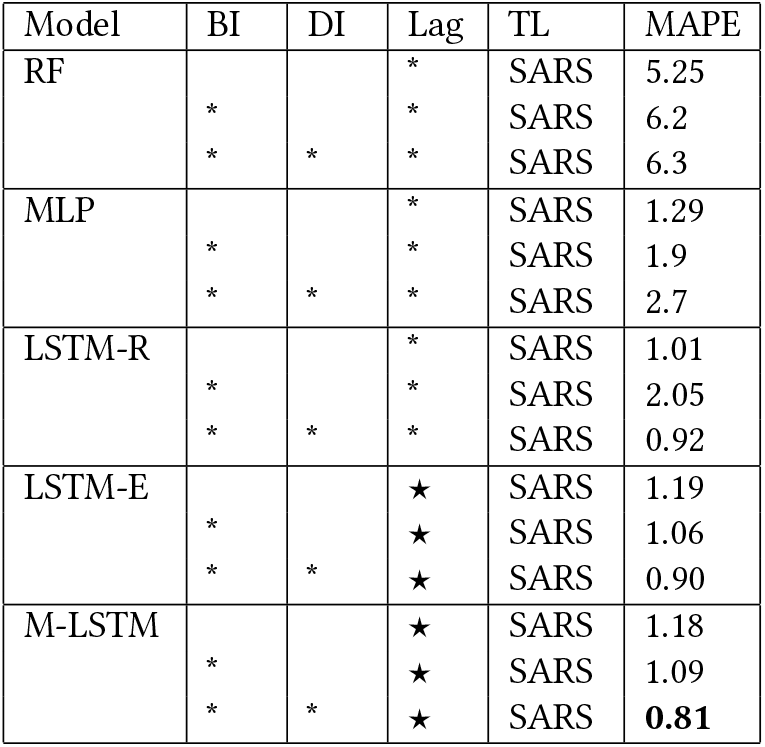
Comparison of the performance of different models for confirmed group. * represents only confirmed cases and ★ shows the confirmed cases along with death and recovered cases. BI stands for basic information, DI for detailed information and TL for transfer learning

The performance of each model is evaluated on the test set of to provide a fair evaluation based on the MAPE value. The best Lag is found by comparing MAPE values as discussed in 5.1. The results also changed by feeding different input features to each candidate model (elaborated in table 2) in section 5.2. The best model/input combination is then found to forecast for next days in section 5.3.

### 5.1 Optimal lag parameter

The proposed models are tested utilizing time series data types. Different time intervals, termed as “lag”, can be considered before the prediction date to feed the occurrence data into the model. Namely, with a sample lag equal to *l* = 4, the model predicts the occurrence in 10st March is using input values in from l prior data points (in 9th, 8th, 7th and 6th March). Figure 7 is designed to show MAPE value for predicting occurrences of confirmed, death, and recovered cases from COVID-19 when **lags of 1-20 days** are used on validation data in preparatory model to find the optimum lag. The lowest MAPE is found for lags of 6, 8, and 10 days, 5, 6, and 7 days, and 5, 6, and 18 days for confirmed, death, and recovered cases, correspondingly. Therefore, **a lag of six days is found to be the “optimal lag parameter”**.

**Figure 7:**
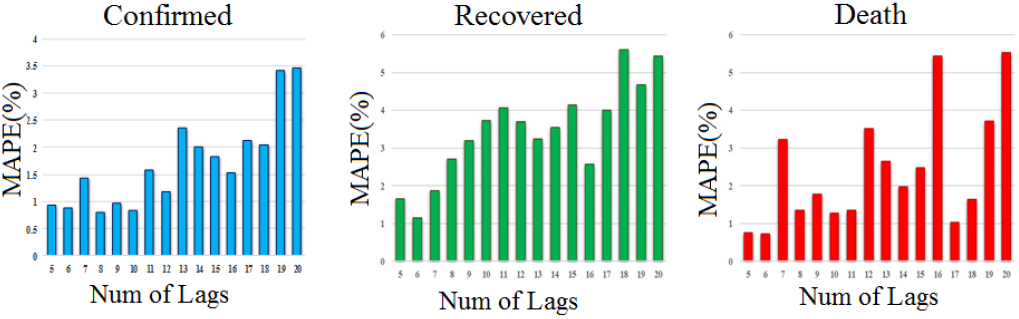
The MAPE values for predicting occurrences of confirmed, death, and recovered cases from COVID-19, when 1-20 days of lag are used in preparatory model.

### 5.2 Optimal Model Selection

Three different types of models are chosen as candidates for this application as elaborated in section 4. To evaluate the performance of the models, it is important to consider different inputs fed into each model. For example, discussing about RF model, three scenarios can be assumed: using basic and detailed features, selecting lag (previous occurrences), and combining all features and lags. This can also be repeated for each model and the performances can be then evaluated.

We selected the MAPE metric to compare different model/ input combinations. The MAPE metric provides the percentage of error on real value and is more reliable compared to Mean Absolute Error which only demonstrates the value of the error (difference of the predicted and real values).

For such an evaluation, a set of nine countries are selected; China is undoubtedly the main candidate as the starting point of COVID-19. Iran, Italy, Spain, and USA are selected due to the report of high number of confirmed and death cases. Germany and Switzerland are also coming from different trend with high number of confirmed cases and controlled number of death. Finally, Korea and Japan are also included for demonstrating the countries with high degree of control on the epidemic. Table 2 shows a summary of the performance on nine selected countries with different models and diverse combination of input features.

With respect to results of Table 2, we found a new set of parameters including basic and detailed feature plus lag (previous occurrences). Furthermore, the results suggest that M-LSTM is the best performing network for identifying the true magnitude of the pandemic with a MAPE value of 0.81% for nine selected countries. This network, as elaborated in section 4, uses lag information from confirmed, death, and recovered cases to predict each next occurrence. This result shows that considering the mutual effect of these three occurrences can provide a better modeling and ignoring such dependence, leads to less performance. Table 2 also shows that lowest performance is obtained with RF. Figure 8 compares the ability of best and worst performing models in correct prediction of the test values.

**Figure 8:**
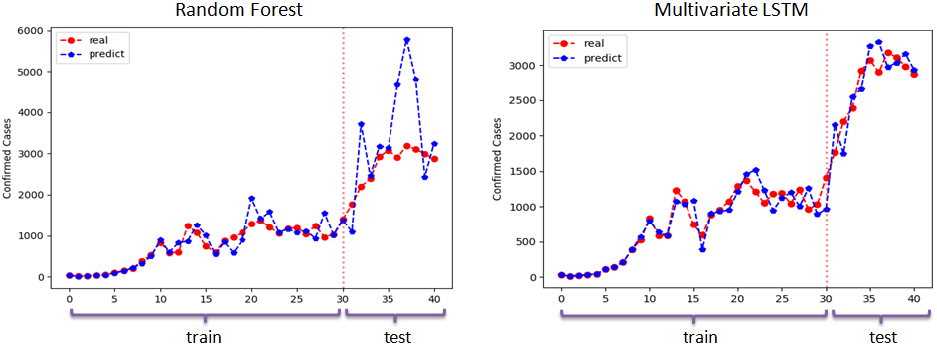
Comparison of the worst and the best performing models in correct following of the training set and accurate prediction of the test values.

### 5.3 Future trajectory of COVID-19 in Iran

To provide a forecasting on number of confirmed, death, and recovered cases in Iran, we predict the upcoming days until July 12nd of 2020. Considering results in Table 2, the best performing network (M-LSTM) is selected with the best combination of the inputs (Basic information, detailed information and lags from all three kind of occurrences). The forecasting is illustrated in figure 9. Figure 9 a and b show the predicted daily and cumulative number of confirmed, death, and recovered cases in Iran. The performance of the forecasting is also presented after April 2 and the values are compared to real reported values in Figure 9 c. To show the performance of the model on other countries, We also present the predictions by proposed method for Japan and Germany in Appendix A.

**Figure 9:**
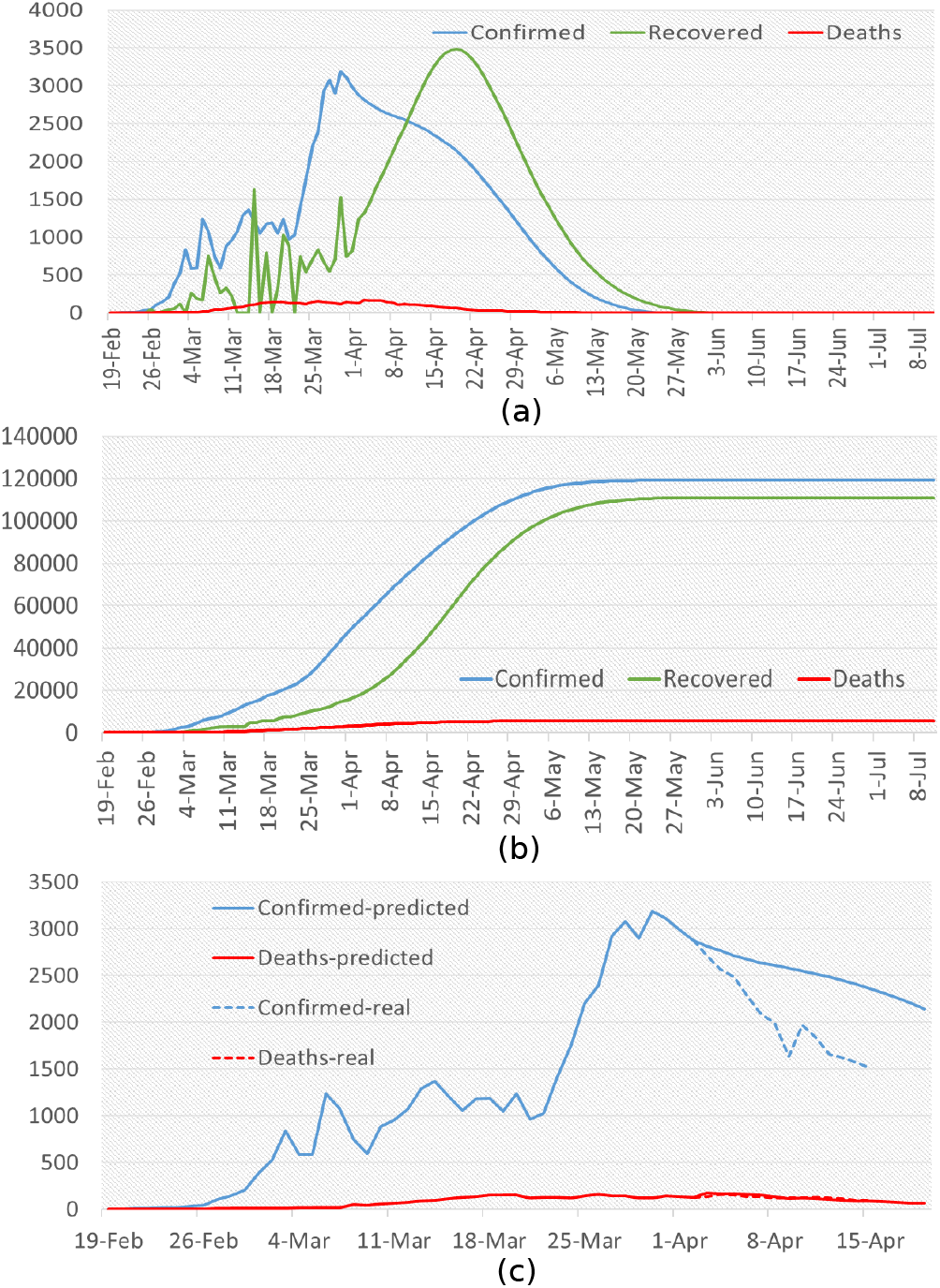
Forecasting confirmed, death, and recovered values for Iran using M-LSTM method for (a) daily and (b) cumulative values, and performance of the forecasting after April 2 compared to real reported values.

#### 5.4 Effect of country actions on predictions

To show how the country actions may change the trend related to the number of infected people and show how older data may reveal the possible scenarios in case of different reactions by countries, we stopped the training process in three different time points and predicted the next-coming days. Three main actions and occasions in Iran are considered in this paper:

- The nationwide closure of schools/universities, nationwide closure of non-essential services and bazaars and closure of metros in big cities before March 11.
- Persian new year in March 19 and unfortunate start of travels (which was not banned officially and caused a great amount of transfer in Iran)
- Closure of roads between cities from March 27 to April 4 by the police.

As it can be seen in Figure 10, the training of the model (for predcition of confirmed cases) is stopped in three dates (date March 11, March 23 and April 2 related to three above mentioned occasions). The blue curve in March 11 indicates that without closure of schools/universities and non-essential services, the curve could raise in March 11th. On the other hand, the red curve shows a considerably lower peak could potentially happen if the travelings would not happen due to start of holidays in Iran. Finally, the green curve shows that closure of roads could reduce the number of affected people but since the curve was in downward route, its effect is not a raise in the predicted numbers.

**Figure 10:**
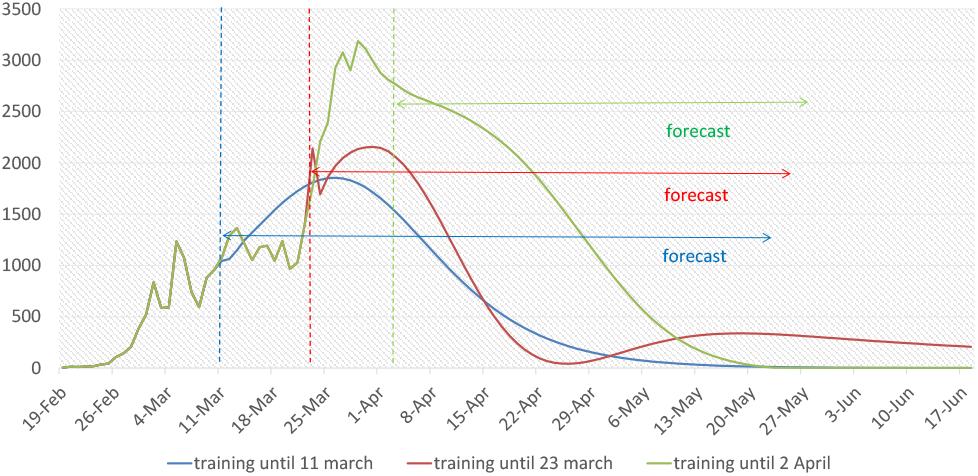
Effect of country actions on predictions. The training on confirmed cases is stopped in three dates (date March 11, March 23 and April 2 related to three main decisions of the country). The curves after mentioned dates, shows what could happen without such decisions.

**Figure 11:**
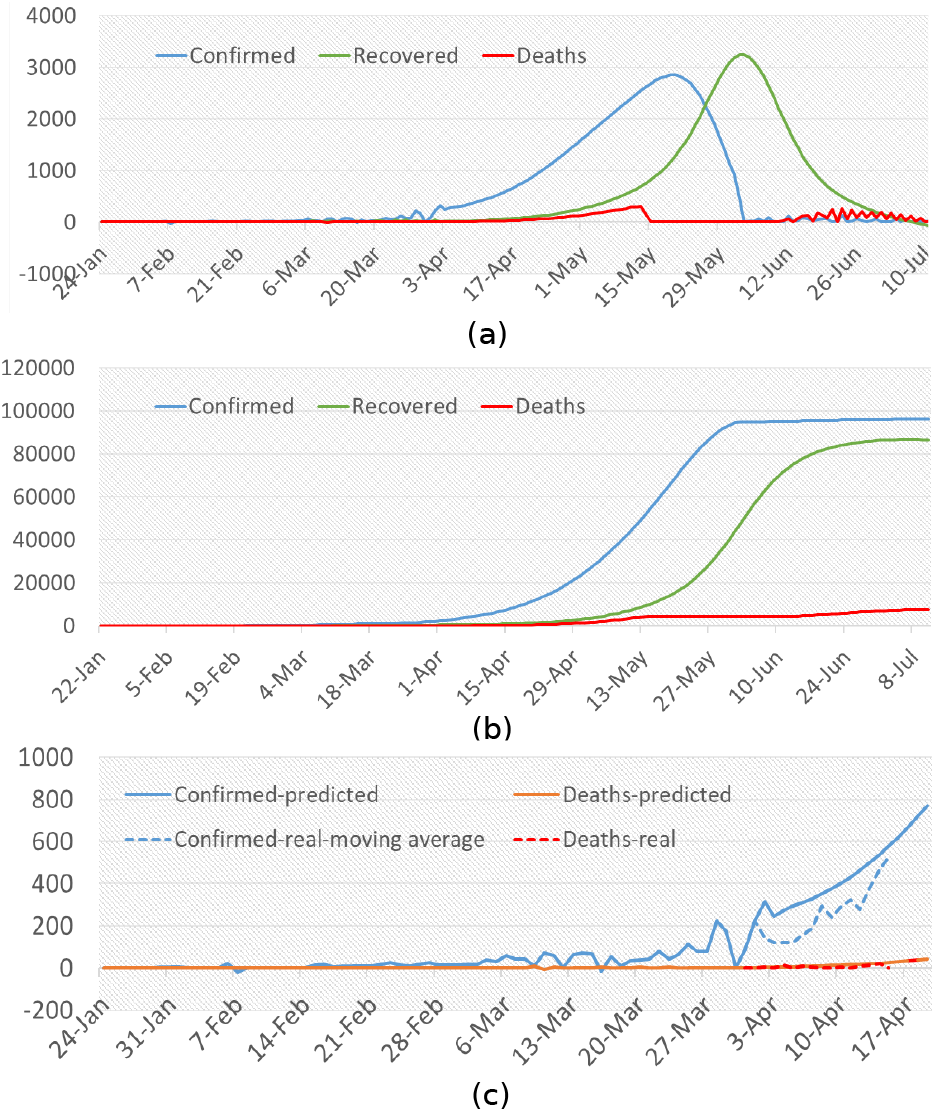
Forecasting confirmed, death, and recovered values for Japan using M-LSTM method for (a) daily and (b) cumulative values, and performance of the forecasting after April 2 compared to real reported values.

**Figure 12:**
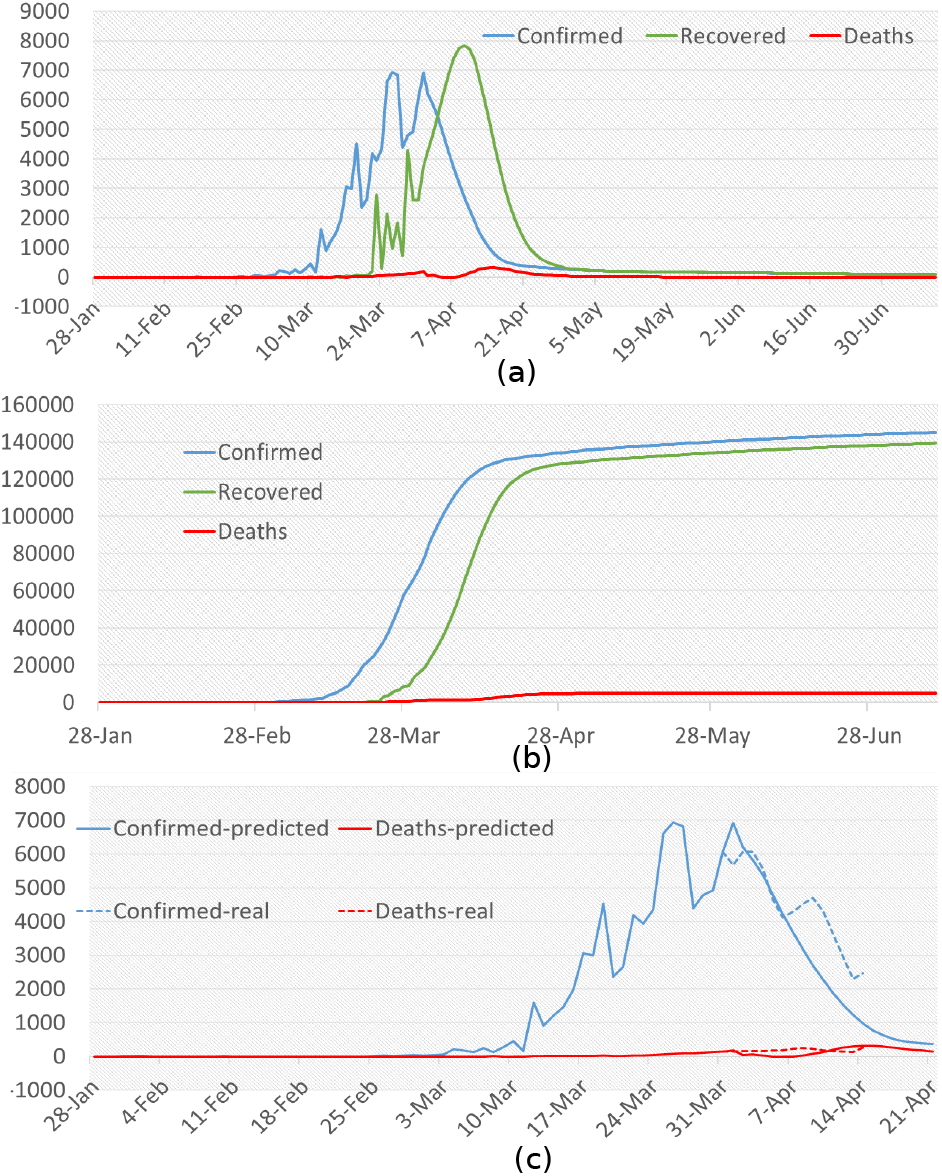
Forecasting confirmed, death, and recovered values for Germany using M-LSTM method for (a) daily and (b) cumulative values, and performance of the forecasting after April 2 compared to real reported values.

## 6 DISCUSSION

CoVid-19 pneumonia started in late December 2019 and pose a continuing and dynamic threat globally. The first case was confirmed by February 19th in Iran. The main question of public and also politicians is the behavior of the epidemic including the peak day, peak number, end point and also daily number of new cases and death. Being aware of real time behavior of epidemics is vital for efficient logistics in the outbreak response. Forecast models will help the policy makers to speculate potential trajectory of the outbreaks and drive interventions as well as estimate the impact of interventions. Several studies has promised efficient tools for forecasting infectious disease dynamics, but these tools are not completely responsible to critical public health needs or have not been evaluated on experimental data.

Different models were tested and based on the minimum Mean Absolute Percentage Error (MAPE) which is an index for accuracy, M-LSTM was the most accurate model found in this study. Using this model we aimed to identify 1) the intensity of the epidemic peak, 2) the timing of the peak, and 3) the total number of cases expected over the duration of the epidemic of COVID-19 in Iran. Determining these outcomes could improve the allocation of resources for risk communication, primary prevention, secondary prevention and preparedness plans (e.g., planning medical staffs, preparing triage units, etc.). In this model we have used the best combination of inputs including lag, basic and detailed information features. In order to have a realistic prediction, three parameters of number of infected cases, number of deaths and number of recovered cases were used together to better shape the epidemic curve. Our simulation model is trained based on publicly available data from all countries of the world and also official reports of the Iranian Ministry of Health and Medical Education from 22 January, 2020 till 2 April 2020.

As illustrated in fig 9 by considering the stability of outbreak response, the peak of the epidemic has already occurred around April 1st, with about 3000 new cases and the epidemic would be ended by early-may with a MAPE of 0.8 percent. The basic assumption of the models is stability of the environment measurements but as we do not live in controlled conditions, every decision would change the epidemic track. The effect of governmental decisions and public occasions is illustrated in figure 10. Considering that it takes around 5-6 days (the median of incubation period) for the results of interventions to show up on new case numbers, the difference between blue and green line in between March 11th and 25th is the result of decisions on school and university closure and cancellation of cultural and religious events. Also the difference between red and green line between March 23rd and April 2nd is because of New Year holiday and travels. It means that by focusing on social distancing there would be a steady decline in new cases after April 2nd as shown by green line but changes in public health policies may change the epidemic track and postpone the end point as well.

Our result could be useful in preparing for future outbreaks as well as current one by considering the results in public health decision making. Similar to other modeling technique, the approach presented here is subject to limitations, which include data quality associated with real-time modeling (as data is often subject to ongoing cleaning, correction, and reclassification of onset dates as further data become available), reporting delays, and problems related to missing data.

## Data Availability

This study uses four data sources to predict COVID-19 disease, including COVID-19 data, basic information, detailed information for
each country, as well as SARS Data. The COVID-19 Data (by John
Hopkins university) contains daily number of confirmed/death/recovered people. The Basic information contains the information about date/country/province of the cases. The detailed
information for each country (around 18576 observations) includes information such as Region/ Population/Area (sq. mi.)/ Pop.
Density (per sq. mi.)/ Coastline(coast/area ratio)/ Net migration/
Infant mortality (per 1000 births)/GDP/Literacy (%)/ Phones (per
1000)/ Climate/ Birthrate/ Death-rate/ Agriculture/ Industry/ Service/Arable (%)/ Crops(%)/. The Occurrences of SARS Data (2539
observations) contains information on the number of confrmed/death/recovered people for SARS.

https://coronavirus.jhu.edu/map.html

https://data.humdata.org/dataset/novel-coronavirus-2019-ncov-cases

## A PREDICTIONS ON MORE COUNTRIES

Here we present the predictions by the proposed model for Japan and Germany in figure and. As it can be seen from figure A (c) and 12 (c), here is always a lag between the actual numbers and the predicted ones but model seems to work on a wide range of countries which is encouraging.

